# Protecting against Respiratory Syncytial Virus: An online questionnaire study exploring UK parents’ acceptability of vaccination in pregnancy or monoclonal antibody administration for infants

**DOI:** 10.1101/2024.05.28.24308012

**Authors:** Simone Paulson, Alasdair PS Munro, Katrina Cathie, Helen Bedford, Christine E Jones

**Affiliations:** NIHR Southampton Clinical Research Facility and Biomedical Research Centre, University Hospital Southampton NHS Foundation Trust, Southampton, UK; Faculty of Medicine and Institute for Life Sciences, University of Southampton, Southampton, UK; Great Ormond Street Institute of Child Health, University College London, London, UK

## Abstract

**Introduction:** Maternal vaccination and infant monoclonal antibodies are promising avenues to protect young infants from respiratory syncytial virus (RSV) infection. Successful inclusion into the UK immunisation schedule depends on parental acceptability, among other factors.

**Methods:** An online cross-sectional survey from August to September 2023 exploring the likelihood of accepting, and preference for, either method of RSV prophylaxis, and reasons given for these. A questionnaire was distributed via social media networks to UK participants with a child under the age of 2 years and/or pregnant.

**Results:** A total of 1620 participants completed the survey. Participants’ median age was 33 years (IQR 31 −36), 92% were of White ethnicity. Acceptability was high, but higher for a maternal vaccine than an infant monoclonal antibody (p<0.0001). Concerns about safety, need for more information, and number of vaccines given to infants already were common reasons for hesitancy. Lacking knowledge about RSV was associated with a lower likelihood of accepting either option (maternal vaccine: OR 0.32, 95% CI 0.16-0.68, p=0.002; infant monoclonal antibody: OR 0.35, 95% CI 0.19-0.68, p= 0.002), as was identifying as Black, Black British, African or Caribbean ethnic group, or having declined the routinely recommended antenatal vaccines.

**Conclusions:** Whilst most parents would accept a maternal vaccine or infant monoclonal antibody to protect their infant against RSV, understanding preferences, influencing factors and concerns is essential to optimise immunisation programmes. This study highlights the information parents require to make an informed choice about RSV protection.

## Introduction

Respiratory syncytial virus (RSV) is the most common cause of lower respiratory tract infection (LRTI) leading to hospitalisation in children under the age of five years (1). Over a third of RSV hospital admissions occur in the 0-6 months age group (2), and identifying effective prevention strategies for RSV in these infants is an international priority (3). The two most promising avenues for protection are vaccination in pregnancy and passive infant immunisation using a monoclonal antibody (4).

Nirsevimab, a long-acting monoclonal antibody, was recently licensed by the Medicines and Healthcare products Regulatory Agency (MHRA) for use in the UK to prevent LRTI caused by RSV for all children entering their first RSV season (5), the Harmonie, phase 3 trial, estimating effectiveness of 83.2% in preventing RSV LRTI (6). A number of maternal RSV vaccines are also currently in development (7). The recently published phase 3 MATISSE study reported over 80% efficacy against severe medically attended LRTI due to RSV in infants up to 3 months of age (8), with regulatory approval granted in the US, EU and UK (9–11).

The success of any immunisation programme depends on factors that include its acceptability to parents and their willingness to have themselves or their children immunised. Previous studies explored public knowledge of RSV as well as general attitudes to vaccination in pregnancy (12–14) and there is some understanding about the acceptability of monoclonal antibodies for passive immunisation (15). Moreover, high uptake (84%) of Nirsevimab in Luxembourg during the winter of 2023 has been reported (16). However, to our knowledge, there are no published studies exploring the likely uptake if either a vaccine in pregnancy or an infant monoclonal antibody was recommended for routine use by the NHS, or comparing the acceptability of the different methods.

This study aimed to explore parents’ views regarding the different methods of protecting infants against RSV. Understanding factors associated with acceptance or hesitance is essential to implement a successful vaccine programme.

## Methods

A cross-sectional observational study was conducted using an online questionnaire between August and September 2023. Participants were recruited from across the UK using adverts designed and managed by the digital marketing company ‘Nativve’, displayed across several social media platforms, limited to UK based IP addresses.

### Population

Participants were eligible if they lived in the UK and either had a child aged <2years, and/or they/their partner were currently pregnant. Inclusion criteria were assessed by yes/no screening questions at the start of the questionnaire.

A formal power calculation was not performed for this observational study; all complete responses collected during the study period were included.

### Questionnaire

The questionnaire was designed specifically for this study, following a review of the literature and input from a public and patient involvement group of parents of children attending outpatient appointments. A summary of all questions is available in the supplementary appendix.

Prior to starting the questionnaire participants were presented with a combined participant information sheet and consent form (supplementary appendix). Confirmation that the participant had read the information, was aged 18 or over and gave their consent to participate was required prior to commencing the questionnaire.

### Ethics

Ethics approval was granted by the University of Southampton Faulty of Medicine Ethics Committee (ERGO number:82762).

### Data Analysis

Statistical analysis was performed using R studio (Version 2023.09.0+463). Descriptive statistics (count and percentage) were provided for all variables. Difference in acceptability between a vaccine in pregnancy and infant monoclonal antibody was explored using a paired Mann-Whitney U Test. *P* values <0.05 were considered as statistically significant. To describe factors associated with acceptance of either a maternal vaccine or infant monoclonal antibody, multivariable logistic regression models for both demographics and baseline knowledge of RSV were produced. A participant was considered as being likely to accept if they gave a score of ≥9/10. Logistic regression was also performed to described factors associated with a preference for either a maternal vaccine or infant monoclonal antibody.

## Results

The survey was conducted over a six-week period from 16^th^ August 2023 to 27^th^ September 2023. Of the 1797 eligible participants who started the questionnaire, 1620 completed the full survey (completion rate 90.2 %). Only completed questionnaires were included in the analysis.

Participants’ characteristics are detailed in table 1. A figure of participants geographic distribution can be found in the supplementary appendix (supplementary Figure 2).

**Table 1.** Participant Characteristics.

| Characteristic | N = 1,620* |
| --- | --- |
| Age | Median (IQR);<br>33.0 (31.0, 36.0) |
| **Number of Children** |  |
| Number of Children - 0- but currently pregnant | 87 (5.4%) |
| Number of Children - 1 | 969 (60%) |
| Number of Children - 2 | 446 (28%) |
| Number of Children - 3 | 99 (6.1%) |
| Number of Children - 4 | 14 (0.9%) |
| Number of Children - 5+ | 5 (0.3%) |
| Currently pregnant | 258 (16%) |
| **Ethnicity** |  |
| Ethnicity - Any White ethnic group | 1,486 (92%) |
| Ethnicity - Asian or Asian British | 58 (3.6%) |
| Ethnicity - Black, Black British, Caribbean or African | 20 (1.2%) |
| Ethnicity - Mixed or multiple ethnic groups | 36 (2.2%) |
| Ethnicity - Other ethnic group | 13 (0.8%) |
| Ethnicity - Prefer not to say | 7 (0.4%) |
| **Total household income** |  |
| Total household income - Under £10,000 | 9 (0.6%) |
| Total household income - £10,000- £19,999 | 32 (2.0%) |
| Total household income - £20,000- £29,999 | 58 (3.6%) |
| Total household income - £30,000- £39,999 | 91 (5.6%) |
| Total household income - £40,000- £49,999 | 135 (8.3%) |
| Total household income - £50,000- £59,999 | 215 (13%) |
| Total household income - £60,000- £69,999 | 184 (11%) |
| Total household income - Over £70,000 | 847 (52%) |
| Total household income - Not sure/ Prefer not to say | 49 (3.0%) |
| **Received (or partner received) all routinely recommended vaccines during most recent pregnancy** |  |
| Received (or partner received) all routinely recommended vaccines during most recent pregnancy - Yes | 1,406 (87%) |
| Received (or partner received) all routinely recommended vaccines during most recent pregnancy - No | 206 (13%) |
| Received (or partner received) all routinely recommended vaccines during most recent pregnancy - Not sure/Prefer not to say | 8 (0.5%) |
| **Vaccination status of child(ren)** |  |
| Vaccination status of child(ren) - Up to date/fully vaccinated | 1,487 (97%) |
| Vaccination status of child(ren) - Partially vaccinated | 31 (2.0%) |
| Vaccination status of child(ren) - Unvaccinated | 15 (1.0%) |

### Baseline Knowledge of RSV

Most respondents reported good understanding of flu (87%), however fewer reported a good understanding of RSV (34%), See Table 2. More respondents reported never having heard of (11%) or being unfamiliar with (19%) RSV, compared to conditions caused by RSV – bronchiolitis and pneumonia – or to “flu”.

**Table 2.** Disease Familiarity.

| Familiarity | RSV | Bronchiolitis | Flu | Pneumonia |
| --- | --- | --- | --- | --- |
| I have a good understanding and know the problems it can cause | 555 (34%) | 652 (40%) | 1,411 (87%) | 946 (58%) |
| I've heard of it and know something about it | 589 (36%) | 716 (44%) | 203 (13%) | 649 (40%) |
| I've heard of it, but I'm not sure what it is | 302 (19%) | 212 (13%) | 4 (0.2%) | 23 (1.4%) |
| I've never heard of it | 173 (11%) | 38 (2.3%) | 1 (<0.1%) | 2 (0.1%) |
| I don't know/ I'd prefer not to say | 1 (<0.1%) | 2 (0.1%) | 1 (<0.1%) | 0 (0%) |

More participants reported not knowing how serious RSV is (12.2%) compared to bronchiolitis, pneumonia and flu (6.7%, 0.9% and 0.7% respectively). Amongst respondents who did feel able to gauge severity, pneumonia was perceived as the most severe (Figure 1).

**Figure 1:**
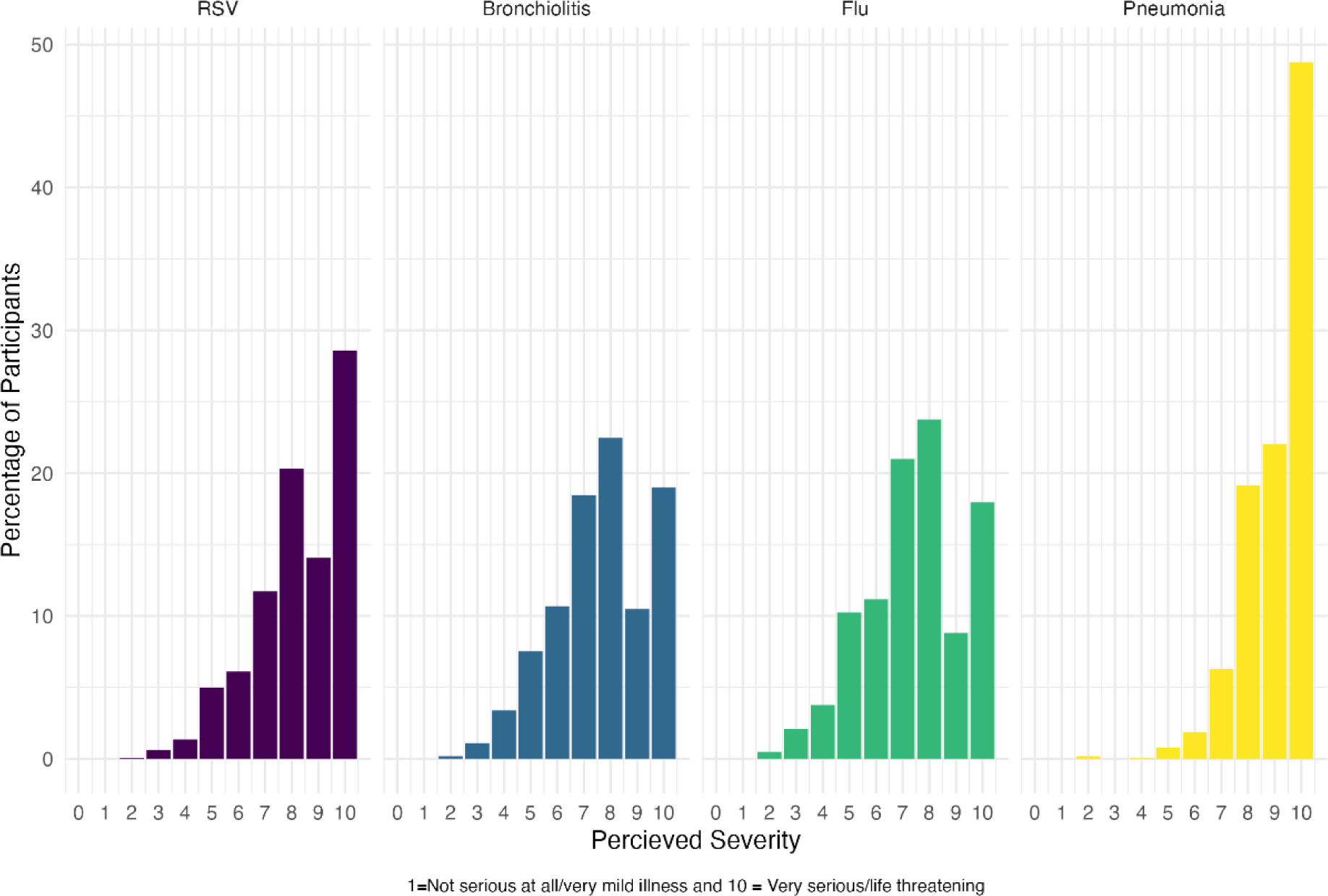
Perceived severity of diseases reported by participants at baseline.

### Acceptability of RSV Vaccine in Pregnancy and Infant Monoclonal Antibody

Acceptability of both a vaccine in pregnancy and infant monoclonal antibody was high, with most respondents reporting that they would be likely (score of ≥9/10) to accept either if routinely recommended for use by the NHS (Table 3, Supplementary Figure 1).

**Table 3.** Vaccine acceptance.

|  | **N = 1,620** |
| --- | --- |
| Maternal Vaccine | 1,422 (88%) |
| Infant Antibody | 1,247 (78%) |
| Infant Antibody- if given before discharge after birth | 1,267 (79%) |

However, acceptability was significantly higher for a vaccine in pregnancy than an infant monoclonal antibody (p<0.0001). Timing of the offer and administration of antibody to the infant either before discharge from hospital following birth or after discharge - and therefore when the infant is a little older - did not affect overall acceptability of antibody (p=0.59).

Most parents (n = 1531, 94.4%) felt that being offered both vitamin K and an infant antibody by injection, after birth but before discharge from hospital, would not affect their decision to accept or reject an RSV antibody for their infant, with the majority (*n* = 1457, 89.9%) reporting that they would be willing for their child to have both, if routinely recommended, prior to discharge (Supplementary information Table 1).

When asked about a preference, the majority of participants (82.6%) expressed a preference for a vaccine in pregnancy over an infant antibody (Figure 2).

**Figure 2:**
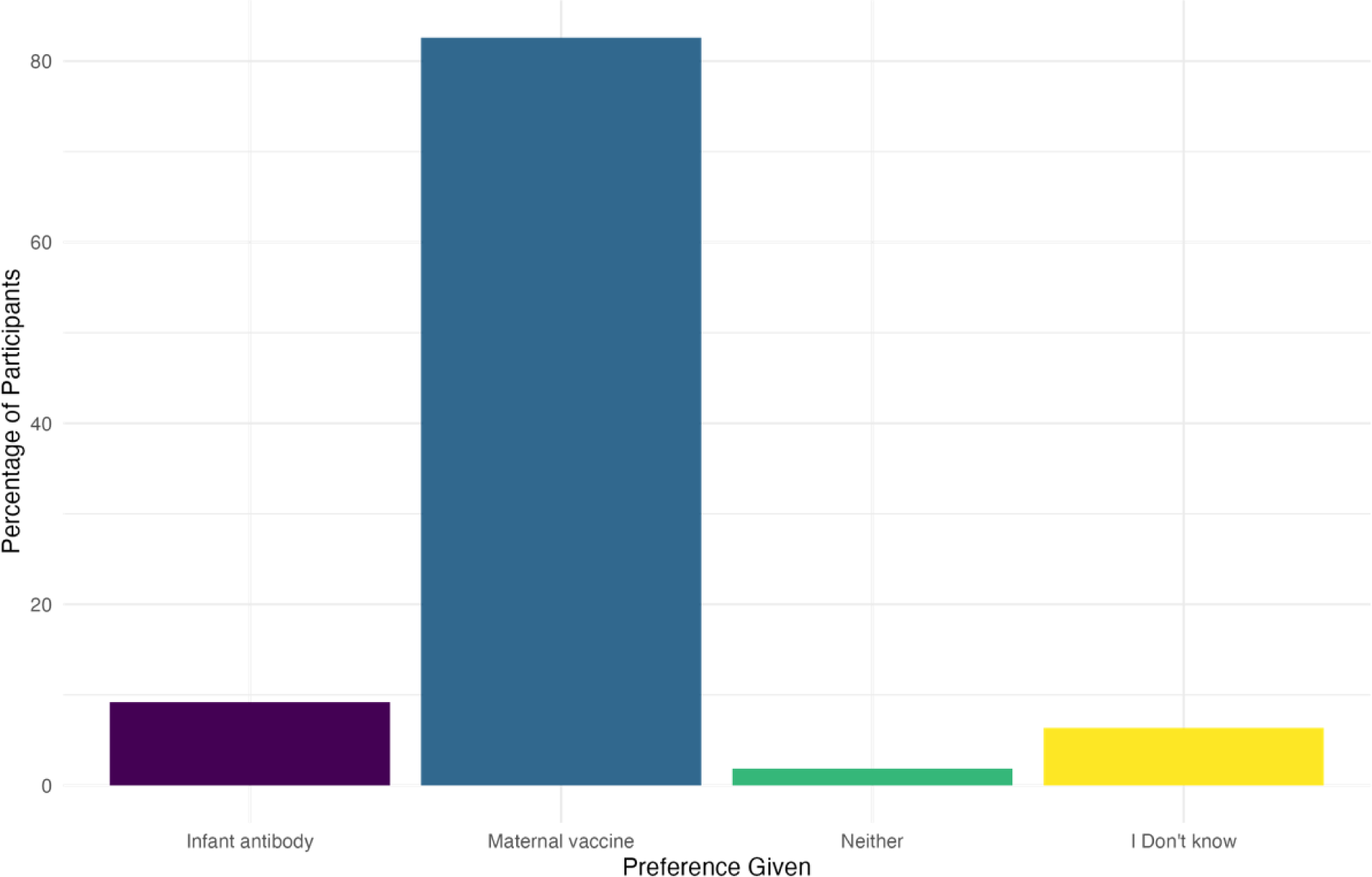
Preference for Vaccine or Antibody.

Desire to protect their infant against RSV, knowledge that the vaccine or antibody was safe and knowledge that the vaccine or antibody was effective were the three most important reasons for accepting an RSV vaccine or infant antibody (Supplementary Table 2). Safety for both the infant and mother, a lack of knowledge about the vaccine and concerns about duration of protection were the most common reasons for hesitancy towards receiving an antenatal RSV vaccine. In addition, concern about the number of vaccines given to infants was a common reason (583/1620, 36% of respondents) for hesitancy about RSV monoclonal antibody administration for an infant.

Safety for the baby remained the most frequently cited reason for hesitancy amongst the subset of participants who would be less likely to receive either a vaccine in pregnancy or an infant antibody (score ≤8/10). This was also the most frequently cited reason by respondents who reported being hesitant about either accepting a vaccine in pregnancy (169/198, 85.4%) or an infant antibody (272/373, 72.7%) (Supplementary Tables 3 and 4.). Not having enough information about the vaccine (103/198, 52.0%) or antibody (205/373, 55%), and concerns about the number of vaccines given both in pregnancy (46/198, 23.2%) and to infants (202/373, 54.2%) were also common reasons for hesitancy.

When asked about reasons for preference for either a vaccine in pregnancy or infant antibody, the most commonly chosen were: ‘I would rather I had the injection than my baby’ (1276/1620, 78.8%), ‘I would accept whichever was recommended by the NHS’ (830/1620, 51.2%) and ‘Number of injections/vaccines for baby’ (502/1620, 31.1%), (Supplementary Table 5).

## Factors Associated with Acceptability of Vaccine in Pregnancy or Infant Monoclonal Antibody

Factors associated with a reduced adjusted OR of scoring ≥9/10 for acceptability of either a vaccine in pregnancy or an infant monoclonal antibody were identifying as being from a Black, Black British, African or Caribbean ethnic group (maternal vaccine: Odds ratio [OR]= 0.09, 95% confidence interval [CI] 0.03-0.3, p<0.001; infant monoclonal antibody: OR=0.19, 95%CI 0.05-0.61, p=0.007) or not having had all the routinely recommended antenatal vaccines (maternal vaccine: OR 0.06, 95%CI 0.04-0.09, p=<0.001, infant monoclonal antibody: OR 0.15, 95%CI 0.10-0.21, p<0.001), (Table 4). Having partially immunised children was associated with lower acceptability for an infant antibody (OR 0.15, 95%CI 0.05–0.39, p<0.001). Having children who had had no routine childhood vaccines was associated with a lower vaccine acceptance in pregnancy (OR 0.21, 95%CI 0.05-0.79, p=0.023), but this did not meet statistical significance threshold if the respondents’ other children were partially vaccinated.

Being less familiar with RSV was associated with lower acceptability of both a vaccine in pregnancy and infant monoclonal antibody (maternal vaccine: OR 0.32, 95%CI 0.16-0.68, p=0.002, infant monoclonal antibody: OR 0.35, 95%CI 0.19-0.68, p= 0.002), Table 4.

Currently being pregnant was significantly associated with a preference for a maternal vaccine (OR 2.2, 95% CI 1.09-4.68, p=0.041), whilst not receiving routinely recommended antenatal vaccines was associated with a preference for an infant antibody (OR 0.4, 95% CI 0.25-0.65, p<0.001) (Table 6).

**Table 4.** Multivariable logistic regression of demographic factors influencing acceptability of RSV immunisation. **Table 4 Legend:** Results indicate adjusted odds of scoring 9 or 10 out of 10 on likelihood of accepting either a maternal vaccine or infant monoclonal antibody as immunisation against RSV.

| Characteristic | Maternal Vaccine |  |  | Infant Antibody |  |  |
| --- | --- | --- | --- | --- | --- | --- |
|  | OR | 95% CI' | p-value | OR | 95% | p-value |
| Age | 1.00 | 0.95, 1.04 | 0.8 | 0.99 | 0.96, 1.02 | 0.5 |
| **How many children do you have?** |  |  |  |  |  |  |
| 1 | - | - | - | - | - | - |
| 2 | 0.63 | 0.42, 0.95 | 0.026 | 0.78 | 0.58, 1.05 | 0.10 |
| 3 | 0.53 | 0.27, 1.11 | 0.082 | 0.81 | 0.48, 1.42 | 0.4 |
| 4 | 0.21 | 0.05, 0.99 | 0.034 | 0.43 | 0.13, 1.63 | 0.2 |
| 5+ | 1.04 | 0.08, 15.3 | >0.9 | 1.80 | 0.18, 20.4 | 0.6 |
| **Pregnant** |  |  |  |  |  |  |
| No | - | - | - | - | - | - |
| Yes | 0.99 | 0.55, 1.84 | >0.9 | 0.84 | 0.56, 1.29 | 0.4 |
| **Ethnicity** |  |  |  |  |  |  |
| Any White ethnic group | - | - | - | - | - | - |
| Asian or Asian British | 0.57 | 0.24, 1.50 | 0.2 | 0.85 | 0.43, 1.80 | 0.7 |
| Black, Black British Caribbean or African | 0.09 | 0.03, 0.30 | <0.001 | 0.19 | 0.05, 0.61 | 0.007 |
| Mixed or multiple ethnic groups | 1.85 | 0.55, 8.63 | 0.4 | 0.85 | 0.38, 2.06 | 0.7 |
| Other ethnic group | 0.94 | 0.13, 23.8 | >0.9 | 8.85 | 0.74, 389 | 0.2 |
| Prefer not to say | 0.33 | 0.05, 3.41 | 0.3 | 0.61 | 0.10, 4.21 | 0.6 |
| **Income** |  |  |  |  |  |  |
| Under £10 000 | - | - | - | - | - | - |
| £10 000- 19 999 | 0.60 | 0.05, 6.43 | 0.7 | 2.34 | 0.34, 16.5 | 0.4 |
| £20 000 £29 999 | 0.79 | 0.07, 7.67 | 0.8 | 3.51 | 0.54, 23.6 | 0.2 |
| £30 000 39 999 | 0.59 | 0.05, 5.15 | 0.6 | 3.41 | 0.54, 22.1 | 0.2 |
| £40 000 49 999 | 0.36 | 0.04, 3.02 | 0.4 | 3.57 | 0.58, 22.7 | 0.2 |
| £50 000- £59999 | 0.52 | 0.05, 4.27 | 0.6 | 3.69 | 0.61, 23.1 | 0.15 |
| £60 000- £69 999 | 0.79 | 0.08, 6.52 | 0.8 | 6.83 | 1.11, 43.7 | 0.036 |
| Over £70 000 | 0.55 | 0.06, 4.36 | 0.6 | 4.36 | 0.73, 26.9 | 0.10 |
| Not sure/Prefer not to say | 0.63 | 0.05, 6.49 | 0.7 | 1.96 | 0.30, 13.4 | 0.5 |
| **Antenatal Vaccines** |  |  |  |  |  |  |
| Yes | - | - | - | - | - | - |
| No | 0.06 | 0.04, 0.09 | <0.001 | 0.15 | 0.10, 0.21 | <0.001 |
| Not sure/Prefer not to say | 0.14 | 0.02, 1.28 | 0.044 | 0.37 | 0.07, 2.19 | 0.2 |
| **Childhood vaccinations** |  |  |  |  |  |  |
| Up to date/fully vaccinated | - | - | - | - | - | - |
| Partially vaccinated | 0.40 | 0.15, 1.05 | 0.066 | 0.15 | 0.05, 0.39 | <0.001 |
| Unvaccinated | 0.21 | 0.05, 0.79 | 0.023 | 0.46 | 0.14, 1.52 | 0.2 |

**Table 5:**
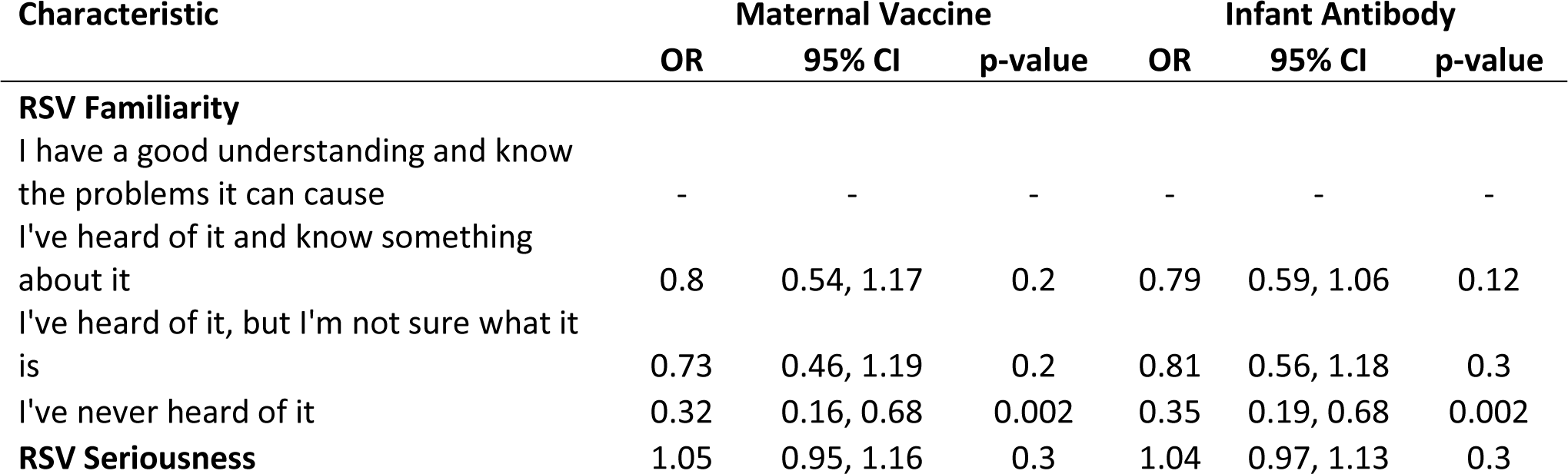
Multivariable logistic regression of baseline RSV knowledge and acceptability of RSV immunisation. **Table 5 Legend**: Results indicate adjusted odds of scoring 9 or 10 out of 10 on likelihood of accepting either a maternal vaccine or infant monoclonal antibody as immunisation against RSV.

**Table 6:** Preference for maternal vaccine or antibody regression model. **Table 6 legend:** Reference is maternal vaccine

| Characteristic | OR* | 95% CI | p-value |
| --- | --- | --- | --- |
| Age | 0.99 | 0.95, 1.04 | 0.8 |
| **How many children do you have?** |  |  |  |
| 1 | - | - | - |
| 2 | 1.29 | 0.85, 1.99 | 0.2 |
| 3 | 0.98 | 0.50, 2.11 | >0.9 |
| 4 | 0.45 | 0.13, 2.12 | 0.2 |
| 5+ | 0.22 | 0.02, 2.46 | 0.2 |
| **Pregnant** |  |  |  |
| No | - | - | - |
| Yes | 2.12 | 1.09, 4.68 | 0.041 |
| **Ethnicity** |  |  |  |
| Any White ethnic group | - | - | - |
| Asian or Asian British | 0.86 | 0.36, 2.56 | 0.8 |
| Black, Black British, Caribbean or African | 0.49 | 0.14, 2.01 | 0.3 |
| Mixed or multiple ethnic groups | 1.72 | 0.50, 10.8 | 0.5 |
| Other ethnic group | 0.56 | 0.13, 4.18 | 0.5 |
| Prefer not to say | 0.54 | 0.07, 11.2 | 0.6 |
| **Income** |  |  |  |
| £10 000 - 19 999 | - | - | - |
| £20 000 - £29 999 | 2.54 | 0.83, 7.76 | 0.1 |
| £30 000 - £39 999 | 5.78 | 1.81, 19.2 | 0.003 |
| £40 000 - £49 999 | 5.82 | 1.96, 17.3 | 0.001 |
| £50 000 - £59999 | 5.91 | 2.10, 16.2 | <0.001 |
| £60 000 - £69 999 | 6.18 | 2.16, 17.4 | <0.001 |
| Not sure/ Prefer not to say | 1.93 | 0.59, 6.47 | 0.3 |
| Over £70 000 | 4.83 | 1.87, 11.8 | <0.001 |
| Under £10 000 | 2.63 | 0.43, 23.9 | 0.3 |
| **Antenatal Vaccines** |  |  |  |
| No | - | - | - |
| Not sure/Prefer not to say | 1.33 | 0.16, 29.4 | 0.8 |
| Yes | 2.49 | 1.54, 3.95 | <0.001 |
| **Childhood vaccinations** |  |  |  |
| No my child(ren) have had some but not all of the routine childhood vaccinations | - | - | - |
| No my child(ren) have not any of the routine childhood vaccinations | 0.83 | 0.10, 9.96 | 0.9 |
| Yes my child(ren) are up to date with the routine childhood vaccinations | 1.12 | 0.28, 3.58 | 0.9 |

The data for acceptability of either a vaccine in pregnancy or infant antibody was highly skewed towards a high-likelihood of acceptance (Supplementary figure 1). Due to this we were unable to fit a linear regression model. However, linear and ordinal models created for comparison and a sensitivity analysis were broadly consistent with the logistic models presented (Supplementary tables 6 - 9).

## Discussion

Our study found high acceptability amongst UK parents for a vaccine administered in pregnancy and monoclonal antibody administered to young infants to protect against RSV. Most respondents expressed a preference for vaccination in pregnancy over a monoclonal antibody administered to young infants, if both were equally safe and effective. Neither timing of administration, nor receipt of both vitamin K and an infant antibody affected the acceptability of a monoclonal antibody. This high acceptability is consistent with previous literature on the acceptability of hypothetical RSV prophylaxis amongst parents in the UK (14, 15). This is the first study to directly compare acceptability of the two most promising methods of protecting infants from RSV, a vaccine in pregnancy or infant monoclonal antibody.

Familiarity with RSV was lower than that of other common respiratory conditions, consistent with previous studies (14, 15). Being less familiar with RSV was associated with reduced odds of acceptance, highlighting the importance of providing appropriate information to accompany the introduction any RSV vaccine or antibody in the immunisation schedule. Interestingly, perceived severity of RSV was not associated with increased odds of acceptance, in contrast to previous literature (17, 18). Our survey was conducted during the summer which could have influenced the results, due to low circulating respiratory viruses during this period.

Participants who self-identified as being from a Black, Black British, Caribbean or African background had significantly lower levels of acceptability of both the maternal vaccine and infant monoclonal antibody, consistent with other studies on ethnographic factors influencing attitudes towards vaccines, albeit the number of respondents identifying as such was low (19, 20). This highlights the importance of addressing specific cultural considerations in preparing resources to help improve uptake of immunisation.

Many parents identified not knowing enough about RSV and the vaccine or antibody as being a barrier to acceptance. Given the likely timing of either administration of a vaccine in pregnancy or monoclonal antibody to newborn infants, information needs to be available and discussed during pregnancy. Midwives will be critical in delivering this information, yet may be less well informed about RSV and prevention. Exploring the knowledge and attitudes of midwives and then developing tailored information and education for midwives, as well as pregnant women and their partners, is crucial.

Acceptability of a vaccine in pregnancy was higher than that for an infant antibody, and many participants expressed a preference for a vaccine administered in pregnancy. The strongest motivator for acceptance of a vaccine in pregnancy or infant monoclonal antibody was a desire to protect the infant. Together, this suggests that emphasising the benefits of vaccination in pregnancy for the infant may help to optimise antenatal vaccine uptake in general.

Preference for a vaccine in pregnancy over infant monoclonal antibody may also reflect an unfamiliarity with monoclonal antibodies. In practice, parents will not be given the option to choose one or the other but offered one option provided within the routine immunisation programme. This data does show that if a monoclonal antibody is recommended, particular attention to the content and delivery of communication will be needed.

To our knowledge this is the largest, and most widely distributed, survey of parents’ attitudes to RSV in the UK to date. Online, untargeted distribution allowed us to achieve a broad geographical representation, including data from all four home nations (Supplementary Figure 2) and a high completion rate.

Our study has several limitations. People with strong views for or against vaccination may be more likely to respond to a survey, making responses less representative. Our sample reported higher uptake of both antenatal and childhood vaccines than current UK uptake of vaccines (87% compared to 60.7% population uptake of antenatal pertussis vaccine in England during 2022-23 (21)). Whilst this may bias our findings towards higher rates of acceptability it is unlikely to have significantly affected the relative acceptability of the two options and preferences expressed.

The sample included more participants self-identifying as being from a white ethnic group (92%) compared to the population as a whole, as well as a higher total household income than the general population (22). In particular, there were a low number of participants reporting as being Black/Black British/Caribbean (20/1620, 1.2%) meaning that findings of this cohort being associated with reduced odds of acceptance of either maternal vaccine or infant monoclonal antibody should be interpreted with caution.

Finally, likelihood of acceptance described by participants here does not necessarily translate into real world uptake. Participants may have felt obliged to say they would accept – a “social desirability” bias. Equally research prior to the introduction of the pneumococcal vaccine estimated acceptability to be 68% (23), yet data following introduction showed much higher rates up of uptake. Data from Luxembourg has been reassuring, with a real world estimated neonatal uptake of 84% in the winter of 2023(16).

Given the importance of safety, careful consideration is required over the communication of the possible increased risk of preterm birth observed in some strata of the trials of the newly approved maternal RSV vaccine. Studies are ongoing to determine what, if any, safety signals exist for this possible complication which led to the previous withdrawal of another candidate maternal RSV vaccine.

## Conclusions

The two most promising methods of protecting healthy term infants from RSV are either a vaccine in pregnancy or an infant monoclonal antibody, but parental acceptability is key to their effectiveness. We have found that, if equally safe and effective, a vaccine in pregnancy would have higher acceptability than an infant monoclonal antibody, but both would have high acceptability. Importantly, acceptability of infant monoclonal antibody is not affected by timing of administration.

This study provides important information for assessing cost-benefit of any newly implemented RSV programme. It also provides further insight into factors associated with likelihood of vaccine acceptance, specific to RSV, essential to optimise the effectiveness of any newly implemented programme. Future work should focus on scoping the informational needs surrounding RSV, how these might differ between subsets of the population, and how to communicate the safety and effectiveness of the proposed intervention.

## Data Availability

Data available on reasonable request

## Author Contributions

The authors made the following contributions to the manuscript. CEJ conceived the original idea and was chief investigator. SP, HB, KC and CEJ designed the questionnaire. SP oversaw data collection, drafted the original manuscript and was principal investigator. SP, APSM and CEJ performed the data analysis. All authors have read and approved the final published version of the manuscript.

## Institutional Review Board Statement

The study was conducted in accordance with the Declaration of Helsinki, and approved by the Faculty of Medicine Ethics Committee, University of Southampton (ERGO number 82762, 14^th^ August 2023).

## Informed Consent Statement

Informed consent was obtained from all subjects involved in the study

## Acknowledgments

The authors would like to thank all the parents who participated in this study, the PPI Team at University Hospital Southampton (Heather Parsons, Rachel Hart, Traci Carroll and Alison Thompson), the team at Nativve Health Research, Dr Helen Moyses and Professor Thomas House for their statistics advice.

## References

1. O’Brien KL, Baggett HC, Brooks WA, Feikin DR, Hammitt LL, Higdon MM, et al. Causes of severe pneumonia requiring hospital admission in children without HIV infection from Africa and Asia: the PERCH multi-country case-control study. The Lancet. 2019;394(10200):757–79.

2. Li Y, Wang X, Blau DM, Caballero MT, Feikin DR, Gill CJ, et al. Global, regional, and national disease burden estimates of acute lower respiratory infections due to respiratory syncytial virus in children younger than 5 years in 2019: a systematic analysis. The Lancet. 2022;399(10340):2047–64.

3. Modjarrad K, Giersing B, Kaslow DC, Smith PG, Moorthy VS. WHO consultation on Respiratory Syncytial Virus Vaccine Development Report from a World Health Organization Meeting held on 23-24 March 2015. Vaccine. 2016;34(2):190–7.

4. Esposito S, Abu Raya B, Baraldi E, Flanagan K, Martinon Torres F, Tsolia M, et al. RSV Prevention in All Infants: Which Is the Most Preferable Strategy? Frontiers in Immunology. 2022;13.

5. MHRA. Final Decision Letter: https://cms.mhra.gov.uk/pip/mhra-100067-pip01-21-m01-update. 2022.

6. Drysdale SB, Cathie K, Flamein F, Knuf M, Collins AM, Hill HC, et al. Nirsevimab for Prevention of Hospitalizations Due to RSV in Infants. New England Journal of Medicine. 2023;389(26):2425–35.

7. PATH. RSV Vaccine and mAb Snapshot | PATH: https://www.path.org/resources/rsv-vaccine-and-mab-snapshot/. 2023.

8. Kampmann B, Madhi SA, Munjal I, Simões EAF, Pahud BA, Llapur C, et al. Bivalent Prefusion F Vaccine in Pregnancy to Prevent RSV Illness in Infants. The New England journal of medicine. 2023.

9. (JCVI) TJCoVaI. Respiratory syncytial virus (RSV) immunisation programme for infants and older adults: JCVI 2023 [Available from: https://www.gov.uk/government/publications/rsv-immunisation-programme-jcvi-advice-7-june-2023/respiratory-syncytial-virus-rsv-immunisation-programme-for-infants-and-older-adults-jcvi-full-statement-11-september-2023#:~:text=Pfizer%20have%20developed%20a%20bivalent,clinical%20trial%20(reference%207).

10. Agency EM. Abrysvo 2023 [Available from: https://www.ema.europa.eu/en/medicines/human/EPAR/abrysvo.

11. (FDA) USFaDA. Abrysvo 2023 [Available from: https://www.fda.gov/vaccines-blood-biologics/abrysvo.

12. Dempsey AF, Pyrzanowski J, Donnelly M, Brewer S, Barnard J, Beaty BL, et al. Acceptability of a hypothetical group B strep vaccine among pregnant and recently delivered women. Vaccine. 2014;32(21):2463–8.

13. Giles ML, Buttery J, Davey MA, Wallace E. Pregnant women’s knowledge and attitude to maternal vaccination including group B streptococcus and respiratory syncytial virus vaccines. Vaccine. 2019;37(44):6743–9.

14. Wilcox CR, Calvert A, Metz J, Kilich E, MacLeod R, Beadon K, et al. Attitudes of Pregnant Women and Healthcare Professionals Toward Clinical Trials and Routine Implementation of Antenatal Vaccination Against Respiratory Syncytial Virus: A Multicenter Questionnaire Study. The Pediatric Infectious Disease Journal. 2019;38(9).

15. Lee Mortensen G, Harrod-Lui K. Expert Review of Vaccines ISSN: (Print) ( Parental knowledge about respiratory syncytial virus (RSV) and attitudes to infant immunization with monoclonal antibodies Parental knowledge about respiratory syncytial virus (RSV) and attitudes to infant immunization with monoclonal antibodies. 2022.

16. Ernst C, Bejko D, Gaasch L, Hannelas E, Kahn I, Pierron C, et al. Impact of nirsevimab prophylaxis on paediatric respiratory syncytial virus (RSV)-related hospitalisations during the initial 2023/24 season in Luxembourg. Eurosurveillance. 2024;29(4):2400033.

17. Díaz Crescitelli ME, Ghirotto L, Sisson H, Sarli L, Artioli G, Bassi MC, et al. A meta-synthesis study of the key elements involved in childhood vaccine hesitancy. Public Health. 2020;180:38–45.

18. Dyda A, King C, Dey A, Leask J, Dunn AG. A systematic review of studies that measure parental vaccine attitudes and beliefs in childhood vaccination. BMC Public Health. 2020;20(1):1253.

19. Davies D, McDougall A, Prophete A, Sivashanmugarajan V, Yoong W. COVID-19 vaccination: patient uptake and attitudes in a multi-ethnic North London maternity unit. Postgrad Med J. 2022;98(1164):750-5.

20. Tiley KS, White JM, Andrews N, Ramsay M, Edelstein M. Inequalities in childhood vaccination timing and completion in London. Vaccine. 2018;36(45):6726–35.

21. UKHSA. Prenatal pertussis vaccination coverage in England from July to September 2022 2023 [Available from: https://www.gov.uk/government/publications/pertussis-immunisation-in-pregnancy-vaccine-coverage-estimates-in-england-october-2013-to-march-2014/prenatal-pertussis-vaccination-coverage-in-england-from-july-to-september-2022#:~:text=pertussis%20vaccine%20coverage%20in%20pregnant,the%202021%20to%202022%20financial.

22. (ONS) OfNS. Average household income, UK: financial year ending 2022 2022 [Available from: https://www.ons.gov.uk/peoplepopulationandcommunity/personalandhouseholdfinances/incomeandwealth/bulletins/householddisposableincomeandinequality/financialyearending2022.

23. Bedford H, Lansley M. More vaccines for children? Parents’ views. Vaccine. 2007;25(45):7818–23.

