## Supplementary appendix for "Protecting against Respiratory Syncytial Virus: An online questionnaire study exploring UK parents’ acceptability of vaccination in pregnancy or monoclonal antibody administration for infants"

**Questionnaire Summary**
Briefly, the questionnaire consisted of eligibility screening questions then five main sections:

1. An exploration of baseline knowledge regarding RSV. It is known that perception of disease severity is an important factor in determining uptake of a vaccine. Knowledge of RSV was compared to that of other common respiratory conditions – bronchiolitis, flu and pneumonia.

Information was then provided about RSV, a hypothetical vaccine in pregnancy and infant monoclonal antibody. As the purpose of this study was to explore any difference in acceptability of a vaccine in pregnancy or infant monoclonal antibody in general and no specific product was described. Therefore, participants were told to presume for purposes of answering the questionnaire, that both were equally safe and effective.

2. Acceptability of a vaccine in pregnancy, scored on a scale of 1 (very unlikely to accept) to 10 (very likely to accept) if routinely recommended by the NHS.

3. Acceptability of an infant long-acting monoclonal antibody. Participants were also asked about acceptability if antibody administration was recommended prior to discharge from hospital, after delivery, as it is possible that at certain times of year this may be recommended.

4. Preference for one or other of the protective options.

4. Demographic information.

Aside from participant age, all questions were multiple choice, it was mandatory to complete each question before being able to progress to the next.

**Supplementary Figure 1- Likelihood of accepting a maternal vaccine or infant monoclonal antibody**

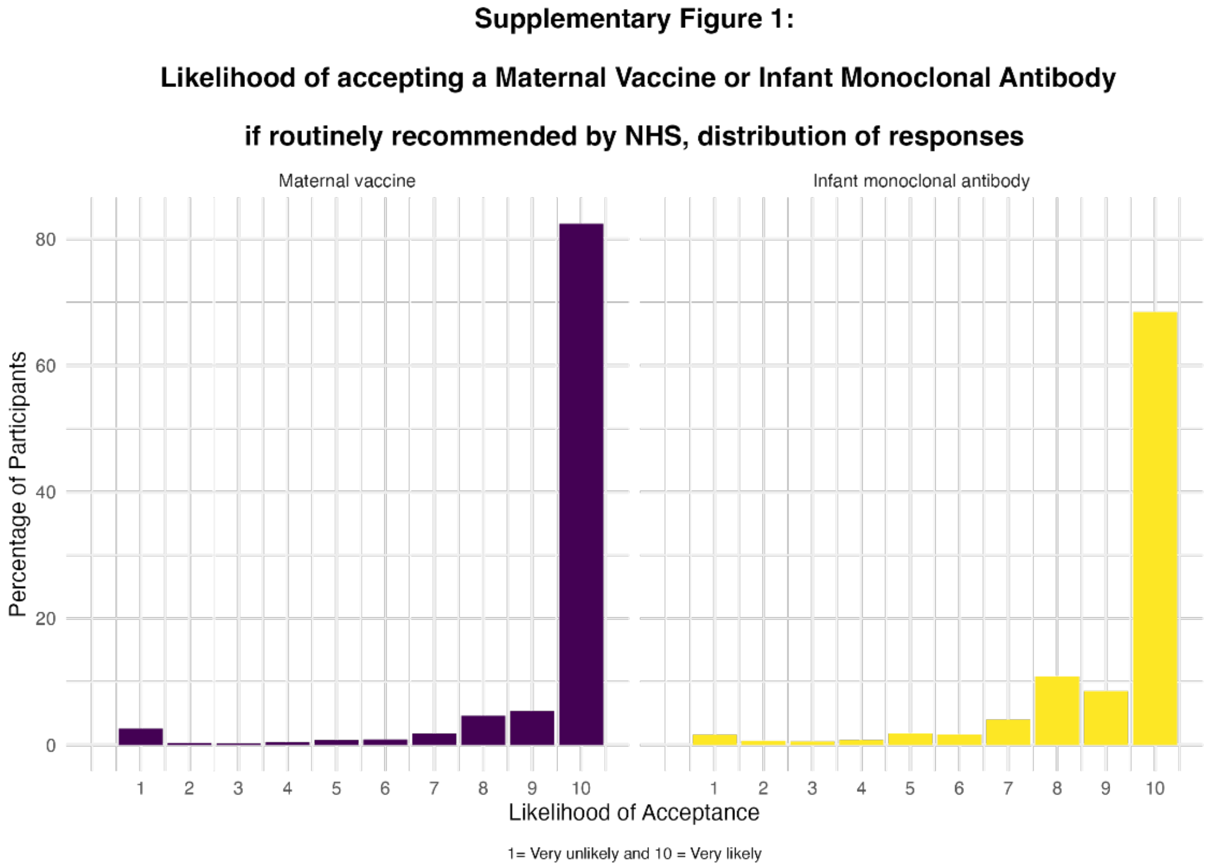

**Supplementary Table 1 - Impact of vitamin K injection prior to discharge on acceptance of infant monoclonal antibody**

**Supplementary Table 1. Would receiving both vitamin K and a RSV antibody by injection, before discharge after delivery, affect your decision regarding accepting an infant antibody for your child?**

| | **N = 1,620**[1] |
|---|---|
| No, I would be happy for my baby to have both before discharge after birth if routinely recommended | 1,457 (90%) |
| Yes, I would want these to be given at different times | 89 (5.5%) |
| No, I would not want my baby to have the antibody before discharge from hospital after birth regardless | 57 (3.5%) |
| I would not want my baby to have vitamin K or the RSV antibody | 17 (1.0%) |

[1] n (%)

**Supplementary Table 2 - Factors influencing decision to accept maternal RSV vaccine**

| What factors would influence you/your pregnant partners' decision to have an RSV vaccine in pregnancy? (Choose up to 3) | Percentage of respondents choosing answer (%) | Count |
|---|---|---|
| To protect my baby against RSV | 86 | 1394 |
| Knowledge that the vaccine is safe | 72 | 1166 |
| Knowledge that the vaccine is effective | 45.4 | 736 |
| Knowledge of RSV and its severity | 34.1 | 552 |
| Recommendation by midwife | 14.2 | 230 |
| Emotional reasons, I would feel guilty if my baby became unwell and I/my partner had not had the vaccine | 11.7 | 189 |
| Avoid any complications of infection | 11.7 | 189 |
| Recommendation by doctor | 8.1 | 131 |
| High likelihood of my child catching RSV | 7.7 | 124 |
| Community/society benefits | 2.5 | 40 |
| Don't know/ other | 0.7 | 12 |
| Recommendation by friends/family | 0.3 | 5 |
| Social pressures | 0.2 | 3 |
| Financial reasons- I can't afford to take time off work if my baby is unwell | 0.2 | 4 |
| Total | | 4775 |
| **What factors would make you/your pregnant partner hesitant to have an RSV vaccine in pregnancy? (Choose up to 3)** | **Percentage of respondents choosing answer (%)** | **Count** |
| I'm worried about the safety of the vaccine for my baby | 56.5 | 916 |
| I don't know enough about the vaccine/I need more information | 34.7 | 562 |
| Duration of protection against RSV | 26.4 | 428 |
| I'm worried about the safety of the vaccine for myself/my partner | 25.6 | 415 |
| Don't know/other | 15.3 | 248 |
| Time/difficulty making appointments for vaccination | 9.4 | 153 |
| Number of vaccines given during pregnancy | 9.2 | 149 |
| Fear of needles | 3.9 | 63 |
| I don't feel that my baby would be at high risk | 3.3 | 53 |
| I prefer natural immunity | 2.8 | 45 |
| I don't feel that RSV is a serious problem | 1.9 | 30 |
| Total | | 3062 |
| **What factors would influence your decision to give your baby the RSV antibody? (Choose up to 3)** | **Percentage of respondents choosing answer (%)** | **Count** |
| To protect my baby against RSV | 86.5 | 1402 |
| Knowledge that the antibody is safe | 72 | 1166 |
| Knowledge that the antibody is effective | 50.6 | 820 |
| Knowledge of RSV and its severity | 29 | 469 |

| | | |
|---|---|---|
| Avoid any complications of infection | 12.5 | 202 |
| Emotional reasons, I would feel guilty if my baby became unwell and I had not given them the antibody | 11.2 | 181 |
| Recommendation by doctor | 10.8 | 175 |
| Recommendation by midwife | 8.7 | 141 |
| High likelihood of my child catching RSV | 8 | 130 |
| Community/society benefits | 2.1 | 34 |
| Don't know/ other | 0.7 | 11 |
| Financial reasons- I can't afford to take time off work if my baby is unwell | 0.3 | 5 |
| Recommendation by friends/family | 0.2 | 3 |
| Social pressures | 0.2 | 4 |
| Total | | 4743 |
| **What factors would make you hesitant to give your baby the RSV antibody?(Choose up to 3)** | **Percentage of respondents choosing answer (%)** | **Count** |
| I'm worried about the safety of the antibody | 51.1 | 828 |
| I don't know enough about the antibody/ I need more information | 39.4 | 639 |
| Number of injections given to babies already | 36 | 583 |
| Duration of protection against RSV | 27.3 | 442 |
| Don't know/other | 15.8 | 256 |
| Time/difficulty making appointments for the antibody injection | 7.7 | 124 |
| I don't feel that my baby would be at high risk | 4 | 65 |
| I prefer natural immunity | 2.3 | 37 |
| I don't feel that RSV is a serious problem | 2.2 | 35 |
| Fear of needles | 1.5 | 25 |
| Total | | 3034 |

**Supplementary Table 3 - Factors contributing to hesitancy towards a vaccine in pregnancy, for those participants who were less likely to accept a vaccine in pregnancy- likelihood score ≤8/10. (n=198)**

| **What factors would make you hesitant to give your baby the RSV antibody? (Choose up to 3) (Participants who are less likely to accept an infant antibody- likelihood score ≤8/10)** | Percentage of respondents choosing answer (%) | Count |
|---|---|---|
| I'm worried about the safety of the vaccine for my baby | 85.4 | 169 |
| I don't know enough about the vaccine/I need more information | 52.0 | 103 |
| I'm worried about the safety of the vaccine for myself/my partner | 42.4 | 84 |
| Number of vaccines given during pregnancy | 23.2 | 46 |
| I prefer natural immunity | 18.7 | 37 |
| Duration of protection against RSV | 16.2 | 32 |
| I don't feel that my baby would be at high risk | 9.1 | 18 |
| I don't feel that RSV is a serious problem | 5.6 | 11 |
| Fear of needles | 4.0 | 8 |
| Don't know/other | 2.0 | 4 |
| Total | | 515 |

**Supplementary Table 4 - Factors contributing to hesitancy towards an RSV antibody, for participants who are less likely to accept an infant antibody- likelihood score ≤8/10. (n=373)**

| **What factors would make you hesitant to give your baby the RSV antibody? (Choose up to 3) (Participants who are less likely to accept an infant antibody- likelihood score ≤8/10)** | Percentage of respondents choosing answer (%) | Count |
|---|---|---|
| I'm worried about the safety of the antibody | 72.7 | 271 |
| I don't know enough about the antibody/ I need more information | 55.0 | 205 |
| Number of injections given to babies already | 54.2 | 202 |
| Duration of protection against RSV | 29.5 | 110 |
| I don't feel that my baby would be at high risk | 9.7 | 36 |
| I prefer natural immunity | 8.6 | 32 |
| Don't know/other | 4.6 | 17 |
| Time/difficulty making appointments for the antibody injection | 4.3 | 16 |
| I don't feel that RSV is a serious problem | 4.0 | 15 |
| Fear of needles | 1.3 | 5 |
| Total | | 909 |

**Supplementary Table 5 - Factors influencing answers to previous questions for those whoare less likely to accept an infant antibody- likelihood score ≤8/10**

| **Which, if any, of the following reasons affected your answer to the previous question?(Preference for a maternal vaccine or infant antibody) (choose up to 4 options)** | Percentage of respondents choosing answer (%) | Count |
|---|---|---|
| I would rather I had the injection than my baby | 78.8 | 1276 |
| I would accept whichever was recommended by the NHS | 51.2 | 830 |
| Number of injections/vaccines for baby | 31 | 502 |
| I’m worried about side effects for my baby | 30 | 486 |
| I think a vaccine in pregnancy would be safer | 19 | 307 |
| I think a vaccine in pregnancy would work better | 8.1 | 132 |
| I don’t know enough about the options/I need more information | 7.7 | 125 |
| I think an antibody for babies would be work better | 6.5 | 106 |
| I’m worried about side effects in pregnancy | 6.3 | 102 |
| I think an antibody for babies would be safer | 4.5 | 73 |
| Other | 3.1 | 50 |
| I prefer natural immunity | 2.2 | 35 |
| Number of vaccines in pregnancy | 1.8 | 29 |
| I don’t feel that RSV is a serious problem | 0.9 | 14 |
| I don’t feel that my baby would be at high risk | 0.7 | 12 |
| Fear of needles | 0.3 | 5 |
| I would be guided by my friends and family | 0.2 | 4 |
| Total | | 4088 |

**Supplementary Table 6 - Multivariable linear regression of demographic factors influencing acceptability of RSV immunisation**

Supplementary Table 6: Multivariable linear regression of demographic factors influencing acceptability of RSV immunisation

| | Maternal Vaccine | | | Infant Antibody | | |
|---|---|---|---|---|---|---|
| **Characteristic** | **Beta** | **95% CI**[1] | **p-value** | **Beta** | **95% CI**[1] | **p-value** |
| **Age** | -0.01 | -0.03, 0.01 | 0.2 | -0.02 | -0.04, 0.00 | **0.041** |
| **Number of Children** | | | | | | |
| 1 | — | — | | — | — | |
| 2 | -0.24 | -0.40, -0.08 | **0.003** | -0.13 | -0.30, 0.05 | 0.2 |
| 3 | -0.25 | -0.55, 0.05 | 0.10 | -0.29 | -0.62, 0.04 | 0.082 |
| 4 | -0.62 | -1.4, 0.12 | 0.10 | -0.34 | -1.2, 0.48 | 0.4 |
| 5+ | 0.26 | -1.0, 1.5 | 0.7 | 0.99 | -0.39, 2.4 | 0.2 |
| **Currently pregnant** | | | | | | |
| No | — | — | | — | — | |
| Yes | -0.11 | -0.33, 0.12 | 0.3 | -0.10 | -0.35, 0.15 | 0.4 |
| **Ethnicity** | | | | | | |
| Any White ethnic group | — | — | | — | — | |
| Asian or Asian British | 0.03 | -0.37, 0.43 | 0.9 | -0.13 | -0.57, 0.31 | 0.6 |
| Black, Black British, Caribbean or African | -2.2 | -2.9, -1.5 | **<0.001** | -1.6 | -2.3, -0.82 | **<0.001** |
| Mixed or multiple ethnic groups | 0.27 | -0.20, 0.74 | 0.3 | -0.04 | -0.56, 0.47 | 0.9 |
| Other ethnic group | 0.38 | -0.43, 1.2 | 0.4 | 0.62 | -0.27, 1.5 | 0.2 |
| Prefer not to say | -1.7 | -2.8, -0.66 | **0.002** | -1.2 | -2.3, 0.01 | 0.052 |
| **Total household income** | | | | | | |
| £10 000- £19 999 | — | — | | — | — | |
| £20 000- £29 999 | -0.14 | -0.76, 0.48 | 0.7 | 0.44 | -0.25, 1.1 | 0.2 |
| £30 000- £39 999 | -0.41 | -0.99, 0.18 | 0.2 | -0.02 | -0.66, 0.63 | >0.9 |
| £40 000- £49 999 | -0.47 | -1.0, 0.08 | 0.10 | 0.02 | -0.60, 0.64 | >0.9 |
| £50 000- £59 999 | -0.22 | -0.76, 0.32 | 0.4 | 0.24 | -0.36, 0.84 | 0.4 |
| £60 000- £69 999 | -0.13 | -0.67, 0.42 | 0.6 | 0.40 | -0.21, 1.0 | 0.2 |
| Not sure/ Prefer not to say | -0.09 | -0.75, 0.56 | 0.8 | -0.02 | -0.74, 0.71 | >0.9 |
| Over £70 000 | -0.19 | -0.71, 0.32 | 0.5 | 0.27 | -0.30, 0.85 | 0.4 |
| Under £10 000 | -0.67 | -1.7, 0.40 | 0.2 | -0.56 | -1.7, 0.61 | 0.3 |
| **Recieved (or partner recieved) all routinely recommended vaccines during most recent pregnancy** | | | | | | |
| No | — | — | | — | — | |
| Not sure/Prefer not to say | 1.0 | -0.11, 2.2 | 0.077 | 1.4 | 0.24, 2.6 | **0.018** |
| Yes | 2.1 | 1.9, 2.4 | **<0.001** | 1.9 | 1.7, 2.1 | **<0.001** |
| **Vaccination status of child(ren)** | | | | | | |
| Partially vaccinated | — | — | | — | — | |
| Unvaccinated | -0.83 | -1.7, 0.06 | 0.067 | 0.24 | -0.73, 1.2 | 0.6 |
| Up to date/fully vaccinated | 2.1 | 1.5, 2.6 | **<0.001** | 2.5 | 2.0, 3.1 | **<0.001** |

[1] CI = Confidence Interval

**Supplementary Table 7 - Multivariable linear regression of baseline RSV knowledge influencing acceptability of RSV immunisation**

| | Beta | 95% CI[1] | p-value | Beta | 95% CI[1] | p-value |
|---|---|---|---|---|---|---|
| **RSV Familiarity** | | | | | | |
| I have a good understanding and know the problems it can cause | — | — | | — | — | |
| I've heard of it and know something about it | -0.11 | -0.31, 0.09 | 0.3 | -0.14 | -0.34, 0.06 | 0.2 |
| I've heard of it, but I'm not sure what it is | -0.05 | -0.31, 0.20 | 0.7 | -0.06 | -0.33, 0.20 | 0.6 |
| I've never heard of it | -0.30 | -0.82, 0.22 | 0.3 | -0.45 | -1.0, 0.09 | 0.10 |
| **RSV Seriousness** | 0.07 | 0.02, 0.13 | **0.009** | 0.09 | 0.03, 0.14 | **0.003** |

[1] CI = Confidence Interval

**Supplementary Table 8 - Multivariable ordinal regression model of demographics on the acceptability of accepting a maternal vaccine or infant antibody**

**Supplementary Table 8: Likelihood of accepting a maternal vaccine or infant antibody, demographics ordinal regression model**

| | Maternal Vaccine | | Infant Antibody | |
|---|---|---|---|---|
| **Characteristic** | **OR**[1] | **95% CI**[1] | **OR**[1] | **95% CI**[1] |
| **Age** | 0.98 | 0.96, 1.01 | 0.97 | 0.94, 1.01 |
| **Number of Children** | | | | |
| 1 | — | — | — | — |
| 2 | 0.73 | 0.57, 0.93 | 0.55 | 0.40, 0.76 |
| 3 | 0.89 | 0.55, 1.46 | 0.77 | 0.43, 1.44 |
| 4 | 0.65 | 0.24, 1.97 | 0.39 | 0.13, 1.41 |
| 5+ | 2.09 | 0.34, 20.1 | 1.16 | 0.15, 13.2 |
| **Currently pregnant** | | | | |
| No | — | — | — | — |
| Yes | 1.02 | 0.71, 1.47 | 0.94 | 0.60, 1.52 |
| **Ethnicity** | | | | |
| Any White ethnic group | — | — | — | — |
| Asian or Asian British | 0.79 | 0.43, 1.48 | 0.80 | 0.38, 1.85 |
| Black, Black British, Caribbean or African | 0.34 | 0.14, 0.81 | 0.17 | 0.06, 0.48 |
| Mixed or multiple ethnic groups | 0.95 | 0.47, 2.08 | 3.07 | 0.98, 13.8 |
| Other ethnic group | 9.00 | 1.26, 209 | 1.96 | 0.28, 53.0 |
| Prefer not to say | 0.48 | 0.11, 2.51 | 0.26 | 0.05, 1.95 |
| **Total household income** | | | | |
| £10 000- £19 999 | — | — | — | — |
| £20 000- £29 999 | 1.88 | 0.75, 4.65 | 1.10 | 0.30, 3.81 |
| £30 000- £39 999 | 1.42 | 0.59, 3.29 | 0.70 | 0.21, 2.12 |
| £40 000- £49 999 | 1.46 | 0.64, 3.21 | 0.68 | 0.21, 1.93 |
| £50 000- £59 999 | 1.79 | 0.80, 3.82 | 0.86 | 0.27, 2.40 |
| £60 000- £69 999 | 2.84 | 1.24, 6.28 | 1.23 | 0.38, 3.53 |
| Not sure/ Prefer not to say | 1.03 | 0.40, 2.58 | 0.90 | 0.24, 3.16 |
| Over £70 000 | 1.83 | 0.85, 3.76 | 1.03 | 0.34, 2.73 |
| Under £10 000 | 0.54 | 0.10, 2.97 | 1.85 | 0.14, 26.1 |
| **Recieved (or partner recieved) all routinely recommended vaccines during most recent pregnancy** | | | | |
| No | — | — | — | — |
| Not sure/Prefer not to say | 2.56 | 0.69, 10.6 | 2.91 | 0.57, 21.6 |
| Yes | 6.91 | 5.03, 9.50 | 13.3 | 9.38, 18.9 |
| **Vaccination status of child(ren)** | | | | |
| Partially vaccinated | — | — | — | — |
| Unvaccinated | 1.32 | 0.39, 4.51 | 0.52 | 0.15, 1.82 |
| Up to date/fully vaccinated | 6.90 | 3.44, 14.0 | 4.76 | 2.17, 10.3 |

[1] OR = Odds Ratio, CI = Confidence Interval

**Supplementary Table 9 - Multivariable ordinal regression of the impact of familiarity of RSV on acceptabilit of maternal vaccine or infant monoclonal antibody**

**Supplementary Table 9: Likelihood of accepting a maternal vaccine or infant antibody, RSV ordinal regression model**

| | Maternal Vaccine | | Infant Antibody | |
|---|---|---|---|---|
| | **OR**[1] | **95% CI**[1] | **OR**[1] | **95% CI**[1] |
| **RSV Familiarity** | | | | |
| I have a good understanding and know the problems it can cause | — | — | — | — |
| I've heard of it and know something about it | 0.82 | 0.59, 1.14 | 0.73 | 0.56, 0.94 |
| I've heard of it, but I'm not sure what it is | 0.61 | 0.41, 0.91 | 0.70 | 0.51, 0.96 |
| I've never heard of it | 0.42 | 0.22, 0.84 | 0.42 | 0.23, 0.76 |
| **RSV Seriousness** | 1.09 | 1.00, 1.18 | 1.07 | 1.00, 1.15 |

[1] OR = Odds Ratio, CI = Confidence Interval

**Supplementary Figure 2 - Distribution of Survey Participants**

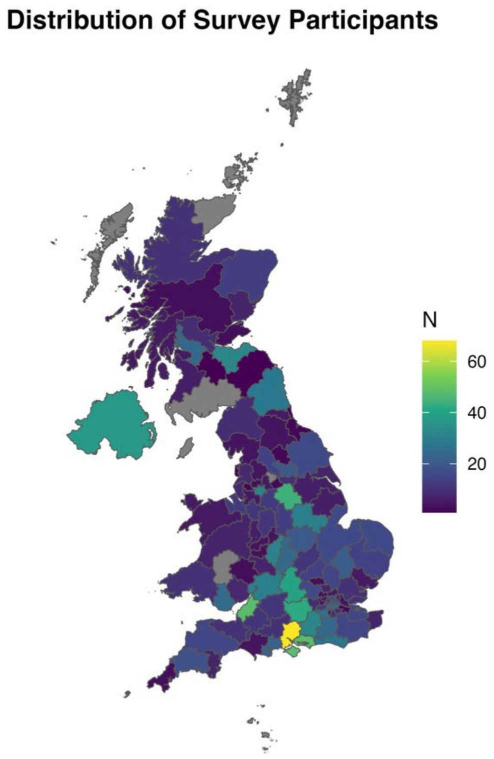

**Participant Information Sheet:**

Introduction:

**Study Title: RSV Protect**

**Researcher(s): Dr Simone Paulson, Dr Katrina Cathie, Dr Helen Bedford, Dr Chrissie Jones**

**University email:** [redacted]

**Ethics/ERGO no: 82762**

**Version: 9 19.07.2023**

You are being invited to take part in a research study by the University of Southampton. Before you continue it is important that you understand why the research is being done and what taking part will involve. Please take the time to read the following information and discuss it with others if you wish.

This study was approved by the Faculty of Medicine Ethics Committee (FOMEC) at the University of Southampton (ERGO Number: 82762).

Why are we doing this study?

This study is to find out what parents think about different ways to protect babies from RSV (respiratory syncytial virus.) Several different ways have been developed, however, currently we don't know how parents would feel about these different options.

It doesn't matter if you have or haven't heard of RSV before, if you decide to take part you will be given some information about this virus.

What happens if I take part?

After you have read this information, you will be asked if you consent to take part. A short, digital survey will then load, this should take no longer than 10 minutes to complete. The questionnaire does not ask for any personal information that could be used to identify you.

At the end of the questionnaire, you will be asked if you would like to be contacted about further studies on this topic. If you would be interested in receiving more information you can fill out your email address. This information will be stored separately to the answers from the questionnaire.

Any contact details shared will be kept strictly confidential and stored in accordance with the University of Southampton's General Data Protection Regulations.

Why have I been asked to participate?

You have been asked to take part because either you have a child under the age of 2 or you/your partner are currently pregnant.
We are aiming to recruit around 1000 participants from across the UK for this study.

What information will be collected?

The questions in this survey ask for information about any prior knowledge of RSV (though it doesn't matter if you've never heard of it), your opinions about new ways to protect babies from RSV and then some demographic information about you. The demographic information includes your age, if you have any children, if you/your children have had vaccinations in the past, your ethnicity, your level of education, your total household income and the first letters of your postcode. This demographic information will help us understand how people from different backgrounds feel about the things in this survey, as well as ensure we have asked a representative group from the population.

Do I have to take part?
No, it is up to you if you decide to take part. Your decision will not affect the medical care you, or any of your family, receive in the future.

What are the possible benefits of taking part?

If you decide to take part in this study, you will not receive any direct benefits; however, your participation will contribute to knowledge in this area of research, which may benefit others in the future.

Are there any risks involved?

It is expected that taking part in this study will not cause you any psychological discomfort and/or distress, however, should you feel uncomfortable you can leave the survey at any time.

What will happen to the information collected?

All information collected for this study will be stored securely on a password protected computer and backed up on a secure server. In addition, all data will be pooled and only compiled into data summaries or summary reports. Only the research team will have access to this information.

For information about how the University of Southampton collects and uses your personal information when you take part in one of our research projects, please see our Privacy Notice: https://www.southampton.ac.uk/assets/sharepoint/intranet/ls/Public/Research%20and%20Integrity%20Privacy%20Notice/Privacy%20Notice%20for%20Research%20Participants.pdf]

The information collected will be analysed and results published in a scientific journal. No personal identifiable information will be used in the analysis or publication of results.

The University of Southampton conducts research to the highest standards of ethics and research integrity. In accordance with our Research Data Management Policy, data will be held for 15 years after the study has finished when it will be securely destroyed.

What happens if there is a problem?

If you are unhappy about any aspect of this study and would like to make a formal complaint, you can contact the Head of Research Integrity and Governance, University of Southampton, on the following contact details: Email: [redacted] phone: [redacted]
Please quote the Ethics/ERGO number above. Please note that by making a complaint you might be no longer anonymous.

More information on your rights as a study participant is available via this link:
https://www.southampton.ac.uk/about/governance/participant-information.page
**Thank you for reading this information sheet and considering taking part in this research.**

Please tick (check) the following boxes as appropriate:
(Note you must confirm that you have read the information, that you are over 18 years of age and that you give your consent in order to take part.)

I have read and understood information on this form
I am aged 18 or over
I consent to take part in this survey
I do not consent to take part in this survey

Note: participants will be taken to the end of the survey unless all of "I have read and understood information on this form", I am aged 18 or over", "I consent to take part in this survey" are selected and "I do not consent to take part in this study" is not selected.

**Social media platforms and survey advertising**

Adverts were designed and managed by the digital marketing company ‘Nativve’. They were displayed across Meta platforms, Google platforms, X (formerly Twitter) and Snapchat. No payment was offered to participants for completion of the survey.

The adverts explained that a survey was being conducted to better understand what parents in the UK think about different ways of protecting babies from RSV. Interested parties were then taken to a study landing page with the PIS.
